# AortaPlot: A Modular End-to-End Protocol for Automated Aortic Imaging Analysis, Visualization, and Standardized Clinical Reporting to Support Aortic Surveillance

**DOI:** 10.64898/2026.01.08.26343732

**Authors:** Santhi Raj Kolamuri, Nicole Aguirre, Devina Chatterjee, Nicholas T. Obey, Sangmita Singh, Uwe Fischer, Eric B. Schneider, Prashanth Vallabhajosyula, Chin Siang Ong

## Abstract

**Background:** Quantitative assessment of aortic morphology from computed tomography (CT) underpins diagnosis, surveillance, and risk stratification in aortic disease. However, existing analysis workflows are often fragmented, algorithm-specific, and limited to discrete vessel segments, constraining reproducibility, longitudinal comparison, and integration of emerging computational methods. The objective of this study was to describe AortaPlot, a modular, end-to-end computational framework designed to standardize continuous aortic quantification and generate anatomically contextualized outputs suitable for clinical interpretation, as well as downstream machine learning and longitudinal modeling.

**Methods:** AortaPlot ingests CT imaging data and performs three-dimensional aortic segmentation using a replaceable segmentation engine. A continuous vascular centerline is computed from the aortic root to the aortic bifurcation and parameterized by arc length. Orthogonal cross-sectional planes are resampled at configurable intervals, and vessel diameter is quantified using a geometry-adaptive Sliding Axis algorithm that identifies the maximum orthogonal diameter independent of cross-sectional shape. Twenty anatomical landmarks are automatically detected and embedded within the measurement framework. The system generates landmark-annotated diameter-versus-length profiles, high-resolution regional analyses, standardized radiomic feature sets, annotated full length aortic visualization and fixed-view multi-angle 3D surface visualizations.

**Results:** Across representative normal and pathological cases, AortaPlot reproducibly reconstructed full-length centerlines and generated anatomically ordered, landmark-annotated diameter profiles spanning the entire aorta. Continuous profiling captured physiologic tapering as well as focal aneurysmal dilation, while region-specific oversampling enabled detailed characterization of localized pathology. Outputs are compiled into standardized, surgeon-oriented reports integrating continuous quantitative profiles, focused segment analysis, anatomically contextualized radiomic features, landmark measurements, annotated aortic visualization and six-view 3D visualizations with fixed rotational spacing.

**Conclusions:** AortaPlot establishes a reproducible, extensible framework for continuous aortic morphology analysis that unifies segmentation, measurement, anatomical context, and visualization within a single pipeline. By standardizing representation across the full vessel length and across time, this protocol reduces analytic variability and produces structured outputs directly amenable to radiomics, machine learning, and longitudinal modeling, providing a scalable foundation for reproducible aortic surveillance and future AI-driven risk stratification.

## Introduction

Computed tomography (CT) is central to the diagnosis, surveillance, and operative planning of thoracic and abdominal aortic disease[1,2]. Clinical assessment of aortic pathology relies heavily on diameter measurements, yet current workflows remain fragmented, operator-dependent, and frequently limited to isolated vessel segments rather than continuous assessment of the aorta from root to bifurcation[3,4]. Variability in segmentation methods, measurement definitions, and reporting formats further limits reproducibility and longitudinal comparison across imaging studies and institutions[5].

Recent advances in medical image segmentation and radiomics have enabled automated extraction of quantitative vascular features from CT imaging, with growing interest in their use for risk stratification and disease monitoring[6,7]. However, many existing tools focus on isolated tasks such as segmentation, measurement, or visualization, without providing an integrated, end-to-end pipeline that produces standardized, clinician-ready outputs. In addition, diameter profiling is often presented without anatomical landmarks, reducing clinical interpretability, and many systems are not designed to accommodate evolving segmentation or measurement algorithms[8].

To address these limitations, we present AortaPlot, a modular, end-to-end protocol for automated aortic analysis that transforms raw CT data into standardized quantitative profiles, radiomic features, and surgeon-oriented visual reports. AortaPlot performs continuous centerline-based diameter assessment from the aortic root to the aortic bifurcation, integrates landmark-annotated two-dimensional profiling with standardized multi-angle three-dimensional (3D) visualization, and is engineered to support interchangeable segmentation and measurement methods. This protocol is designed to support reproducible aortic surveillance, facilitate longitudinal comparison, and enable future integration of emerging computational techniques within a consistent analytical framework.

## Methods

### Study Design and Data Source

This protocol was developed and refined using two complementary datasets comprising computed tomography (CT) scans from diverse clinical sources. These datasets were selected to ensure the pipeline’s robustness across different imaging modalities (CTA and non-contrast CT) and patient populations.

### Dataset 1: Aortic Vessel Tree (AVT) Collection

The AVT dataset[9] includes 56 Computed Tomography Angiography (CTA) scans covering the aortic arch and abdominal aorta, with proximal coverage sufficient for continuous centerline extraction and landmark evaluation. The cohort primarily consists of normal aortic anatomy but includes verified cases of abdominal aortic aneurysm (AAA) and aortic dissection (AD). Ground truth segmentation masks, validated by clinical experts, are provided for benchmarking. The data were obtained from multiple centers and are publicly available under the CC BY 4.0 license.

### Dataset 2: AMOS Multi-Organ Benchmark

The AMOS dataset[10] provides a large-scale benchmark comprising 500 contrast-enhanced CT and 100 MRI scans collected from multi-center clinical environments. Each scan includes voxel-level annotations of 15 abdominal organs, including the aorta. Inclusion of this dataset allows for the assessment of segmentation generalization across heterogeneous acquisition protocols. Data are publicly available under the CC BY 4.0 license.

## Image Processing and Segmentation

### Input Standardization

AortaPlot processes imaging data in NIfTI format. To ensure data integrity, a preprocessing module handles legacy or non-standardized formats (e.g., NRRD) prior to analysis. This module performs two critical corrections: (1) Hounsfield Unit (HU) Calibration, identifying unsigned integer data and applying a -1024 shift to restore the standard signed HU scale (where air ≈ -1000 and water ≈ 0); and (2) Orientation Standardization, converting volumes to the standard RAS (Right-Anterior-Superior) coordinate system using SimpleITK to ensure consistent anatomical alignment. Standard NIfTI inputs are ingested directly into the segmentation pipeline.

### Aortic Segmentation

Three-dimensional volumetric segmentation of the aorta was performed using TotalSegmentator[11] (v2.0), a deep learning framework based on the nnU-Net architecture[12]. The output of this process is a 3D digital ’mask’, a binary map where every voxel (3D pixel) belonging to the aorta is labeled as ’1’ and all other tissues are labeled as ’0,’ effectively isolating the vessel’s volume for analysis. To align computational performance with the time constraints of acute clinical workflows, we implemented a targeted Region of Interest (ROI) subsetting protocol within the model’s standard ‘**total**’ task. Rather than computing the full 117-structure whole-body map, inference was restricted to a minimally sufficient anatomical set comprising the aorta, heart chambers, arteries (brachiocephalic trunk and left subclavian artery) and vertebral landmarks (T12-L3), corresponding to the anatomical span required for abdominal landmark inference. This targeted optimization reduced mean inference time by **92.5%** (decreasing from ∼20 minutes to ∼2 minutes per case on standard GPU hardware) while maintaining full-resolution boundary definition for the relevant vascular structures. The pipeline was engineered in Python (v3.10) utilizing the NiBabel (v5.0), NumPy (v1.24), SciPy (v1.11), and scikit-image (v0.21) libraries.

### Centerline Extraction and Anatomical Ordering

The aortic centerline is derived using a graph-based skeletonization approach designed to ensure anatomical continuity from the aortic root to the aortic bifurcation:

1. *Skeletonization*: The binary aortic mask is reduced to a single-voxel medial axis using morphological thinning (scikit-image).
2. *Path Optimization*: The skeleton is converted into a graph structure (NetworkX). To define the centerline, the algorithm extracts the longest continuous path connecting the vessel’s endpoints. This path is anatomically oriented to run from the aortic root (proximal) to the aortic bifurcation (distal) by using the aortic arch (maximum Z-coordinate) as a reference point.
3. *Root Extension*: To capture the geometry of the aortic root and sinuses of Valsalva, the centerline is theoretically extended from the proximal endpoint using linear regression of the local vessel centroids.
4. *Smoothing*: The raw centerline is smoothed using a B-spline interpolation (SciPy, degree=3) to remove discretization artifacts, yielding a continuous 3D curve parameterized by arc length.

### Anatomical Landmark Detection

Twenty standardized anatomical landmarks are automatically identified along the centerline to provide spatial context. Beyond annotation, these landmarks function as semantic anchors that enable consistent anatomical alignment for longitudinal comparison and downstream quantitative analysis, including machine learning and longitudinal modeling. Landmarks are detected using a hybrid algorithm that combines geometric rules (relative arc length positions) with anatomical reference masks provided by TotalSegmentator (e.g., branching vessels and vertebral levels). When anatomical reference information is unavailable or unreliable, fixed distance offsets derived from commonly used anatomical conventions are applied as approximate reference locations. A complete description of landmark definitions and detection rules is provided in the Supplementary Appendix (Table S1).

### Cross-Sectional Analysis and Diameter Measurement

#### Orthogonal Resampling

Cross-sectional planes are reconstructed at 1.0 mm intervals along the smoothed centerline. For each node, a tangent vector is computed to define the local longitudinal axis, and the 3D volume is resampled onto a 60 × 60 mm plane strictly perpendicular to the vessel flow. The 60 × 60 mm plane size is chosen as larger plane sizes may introduce noise from the adjacent aorta in the thoracic region. However, this is adjustable for aortas with highly abnormal anatomy. This ensures that all subsequent measurements represent true cross-sectional dimensions rather than oblique distortions.

#### “Sliding Axis” Measurement Algorithm

To replicate the “double-oblique” measurement standard of clinical 3D workstations, we implemented a geometry-adaptive Sliding Axis algorithm. Within each orthogonal plane, diameter quantification proceeds in two steps:

1. Major Axis (Axis-1): A virtual caliper rotates in 1-degree increments around the binary aortic cross-section to identify the absolute maximum vessel dimension.
2. Orthogonal Minor Axis (Axis-2): A second caliper, constrained to be strictly perpendicular to the Major Axis, sweeps across the cross-section to locate the maximum orthogonal width.

This two-step sliding approach ensures that asymmetric or irregular morphologies of the aorta are measured at their true widest points, avoiding the underestimation risks inherent in centroid-based or equivalent-circle techniques. By enforcing a consistent orthogonal measurement definition along the centerline, this approach also establishes a stable measurement reference that limits the propagation of geometric bias into downstream analyses.

#### Anatomically Adaptive Quality Assurance and Error Correction

To ensure the clinical reliability of longitudinal diameter profiling, we implemented an Anatomically Adaptive Resampling (AAR) protocol to identify and rectify non-physiological measurement artifacts. Given that automated centerline analysis can be susceptible to geometric errors in regions of extreme tortuosity or vessel pathologies, each measurement was dynamically validated against a local longitudinal median (± 3 contiguous segments). A measurement was flagged as a geometric outlier if it demonstrated a non-physiological deviation (> 30%) from the local vessel trend or failed to meet a minimum detection threshold of 1.0 mm. To resolve these artifacts, the algorithm executed a recursive re-interrogation of the aortic volume. By progressively expanding the sampling kernel, increasing effective slice thickness from a baseline of 10 mm up to 25 mm, the system optimized the surface point density. Each iteration was scored via a morphology-matching objective function; the sampling parameters yielding the highest anatomical concordance with the adjacent vessel segments were automatically selected. This self-correcting framework preserves the nuance of focal pathology, while ensuring a smoothed, clinically actionable diameter profile that eliminates the need for manual expert adjudication.

### Longitudinal Surveillance

To enable precise tracking of aortic disease over time, AortaPlot supports automated longitudinal analysis across serial CT scans[13]. Traditionally, aortic monitoring is limited by discrete, isolated measurements where clinicians manually select 2D slices that may not correspond perfectly between scans. This framework resolves that inconsistency by using automated landmark matching to align every imaging study to a standardized, patient-specific coordinate system. This ensures that measurements are co-registered at the exact same anatomical locations along the vessel across years of follow-up[14].

Beyond diameter, AortaPlot assesses aortic lengthening by calculating the 3D arc length of the centerline between fixed landmarks. Because arc length is computed between fixed anatomical reference points, elongation is captured as a structural remodeling signal that is complementary to, and at least partially independent of, diameter change. Vessel elongation[13,14] is a critical marker of structural remodeling and wall stress that often occurs in parallel with dilatation. Additionally, the pipeline enables the longitudinal tracking of radiomic and morphometric features, including regional volume between landmarks. Monitoring these changes, specifically in the aortoiliac segments, is associated with post-interventional outcomes and provides a more sensitive indicator of the aortic “life-cycle” than diameter alone. By unifying widening, lengthening, and volumetric data into a continuous anatomical history, AortaPlot offers a standardized foundation for determining optimal surgical timing.

### Visualization and Reporting

The pipeline generates a standardized, surgeon-oriented report comprising:

1. *Standardized 3D Visualization:* Six rotationally offset (60°) views of the aortic surface.
2. *Whole-Vessel Diameter Profile*: A continuous diameter-versus-length plot annotated with anatomical landmarks.
3. *Focused Segment Analysis*: High-resolution regional profiling with increased sampling density for targeted anatomical segments.
4. *Longitudinal Surveillance*: Overlaid diameter profiles co-registered across time points using anatomical landmarks.
5. *Landmark Annotated Aorta Visualization:* A landmark annotated aortic visualization along with the corresponding sliding-axis measurements.

For 3D surface visualizations, the minor axis diameter was used for rendering landmark discs. The major axis measure was not used to prevent disc protrusion beyond the aortic surface.

## Results

### AortaPlot End-to-End Protocol and Standardized Outputs

AortaPlot implements a modular, end-to-end workflow for automated aortic imaging analysis that transforms raw computed tomography data into standardized quantitative profiles and visual reports (Figure 1). The protocol integrates automated aortic segmentation, centerline computation from the aortic root to the aortic bifurcation, anatomical landmark detection, and continuous cross-sectional diameter measurement into a unified pipeline. Outputs include a landmark-annotated aortic visualization (Figure 2) and diameter-versus-length profile spanning the entire aorta, standardized multi-angle 3D surface visualizations with displayed landmarks as colored discs of diameter matching the minor axis measure (Figure 3), and structured quantitative summaries compiled into a clinician-oriented report. The modular architecture permits substitution of segmentation and measurement components without altering the core workflow.

**Figure 1.**
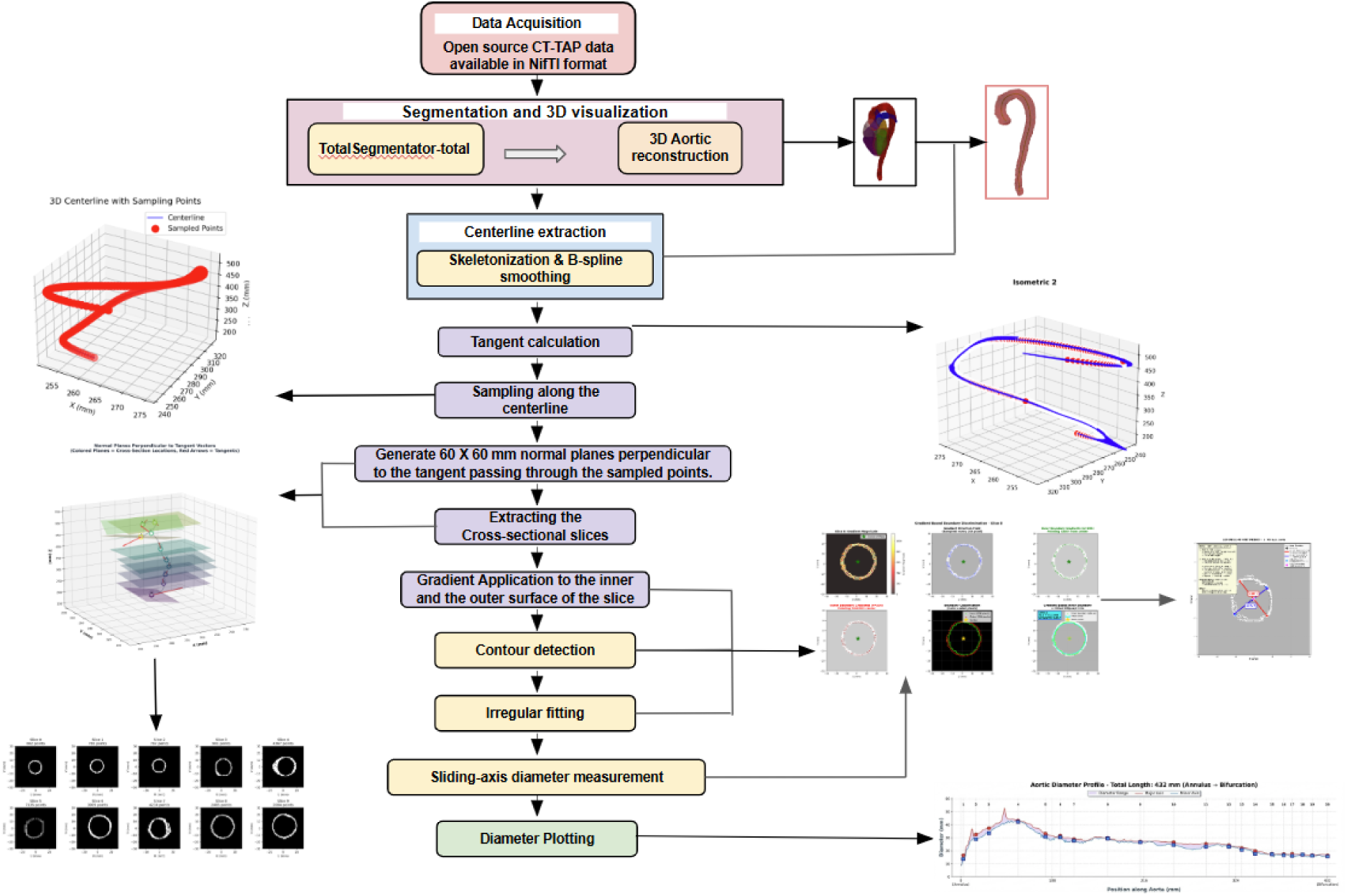
AortaPlot End-to-End Computational Workflow for Continuous Aortic Analysis. Overview of the modular AortaPlot pipeline for automated aortic morphology assessment from computed tomography imaging. Imaging data are ingested in standardized NIfTI format and undergo 3D aortic segmentation using a replaceable segmentation engine. A continuous aortic centerline is extracted from the aortic root to the aortic bifurcation, smoothed, and parameterized by arc length. Orthogonal cross-sectional planes are resampled at fixed intervals along the centerline, and vessel diameter is quantified using a Sliding Axis algorithm that identifies the maximum orthogonal diameter independent of cross-sectional shape. Automated anatomical landmark detection embeds clinical context within the measurement framework. Outputs include landmark-annotated continuous diameter-versus-length profiles, focused high-resolution segment analyses, and standardized multi-angle 3D surface visualizations, which are compiled into a clinician-oriented report.

**Figure 2.**
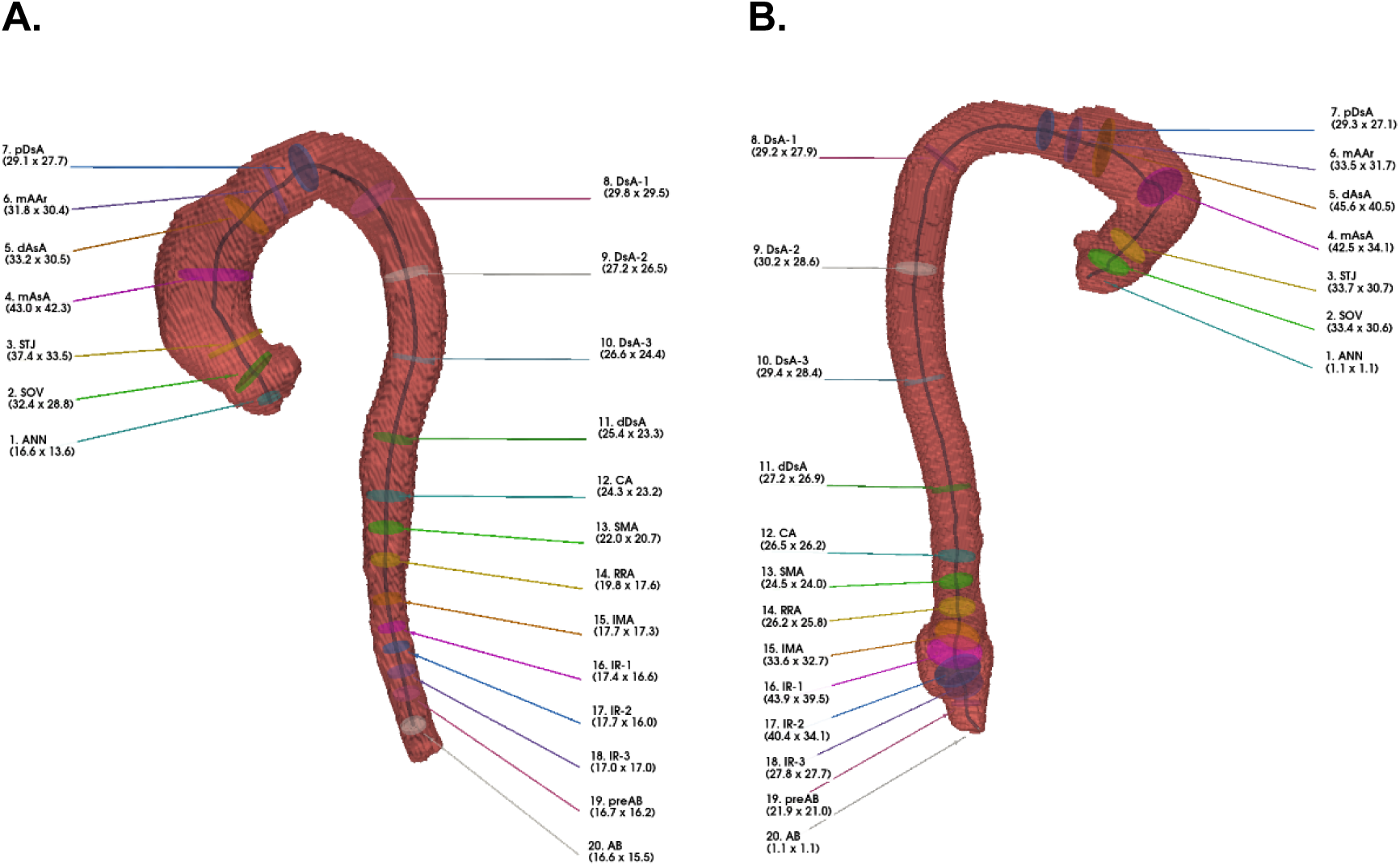
Landmark Annotated Aorta Visualization. Anterior (A) and posterior (B) 3D surface reconstructions of the aorta display comprehensive anatomical landmark annotations. Key regions are identified via centerline-derived analysis and visualized as colored cross-sectional discs. Each call-out label provides a clinical abbreviation (e.g., ANN: Annulus, SOV: Sinuses of Valsalva) and quantitative diameter measurements (Major x Minor axis in mm). This dual-perspective format facilitates the assessment of regional morphology and the identification of focal dilations from the aortic root to the bifurcation.

**Figure 3.**
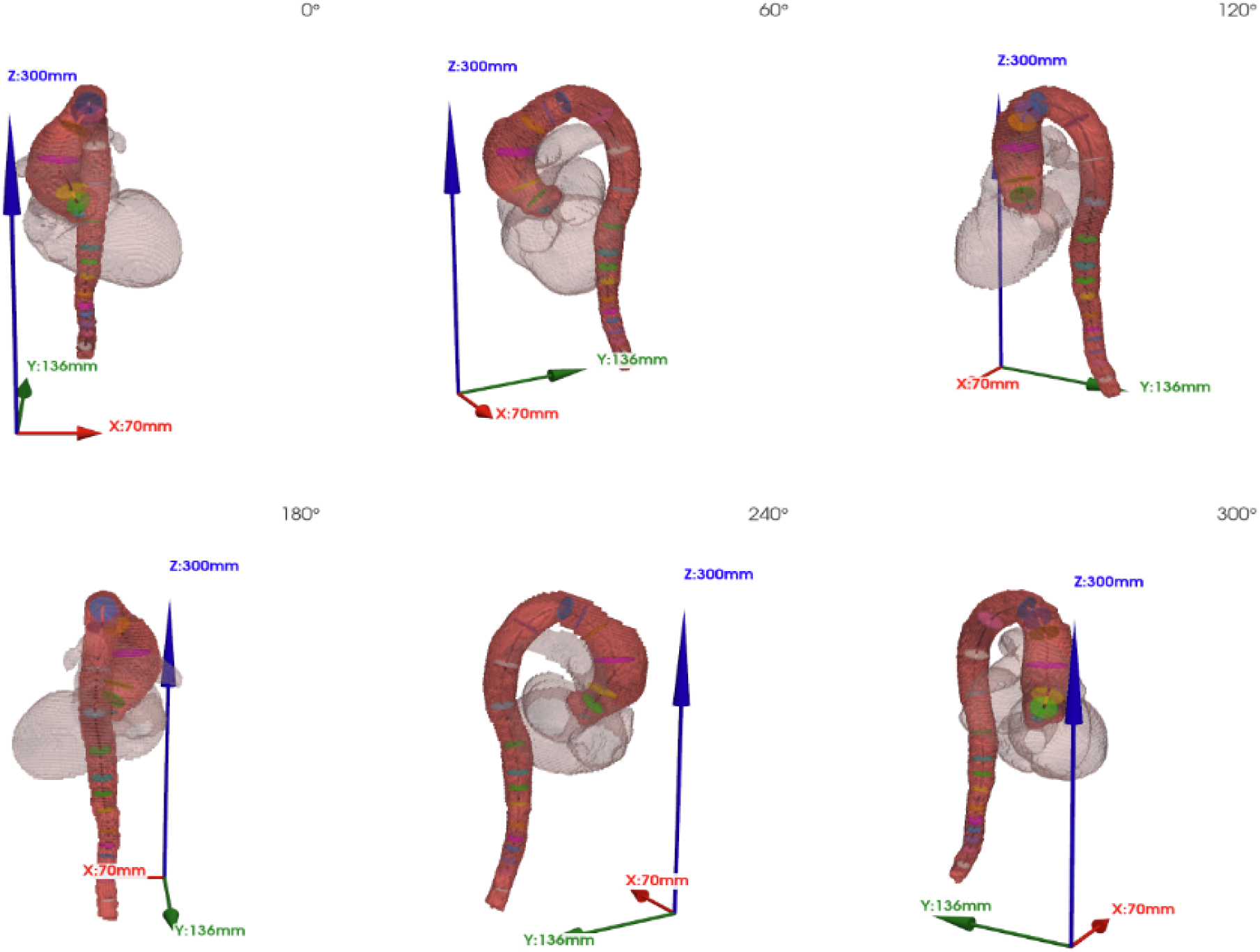
Standardized Multi-Angle 3D Visualization of an Aorta With a Thoracic Aortic Aneurysm Generated by AortaPlot. Six fixed-view 3D surface renderings of the segmented aorta are shown, rotationally offset by 60° increments (0° to 300°) about the longitudinal axis. The standardized viewing geometry enables complete circumferential visualization of aortic morphology within a static report format. Optional semi-transparent cardiac structures provide anatomical context for the ascending aorta and aortic arch. Embedded anatomical landmarks displayed as colored discs using minor axis measurements and centerline-derived reference elements are displayed consistently across views, allowing spatial correlation with quantitative diameter measurements. This multi-angle visualization complements continuous diameter profiling by supporting qualitative assessment of global aortic shape, curvature, and regional dilation.

### Continuous Aortic Profiling in Normal and Pathological Anatomy

Application of AortaPlot to representative cases demonstrates continuous diameter profiling across both normal and pathological aortic anatomy. In a normal aorta, the longitudinal diameter profile shows expected physiological tapering from the ascending thoracic aorta through the abdominal segment, with smooth transitions at annotated anatomical landmarks. In contrast, pathological cases exhibit localized deviations from baseline diameter patterns, readily identifiable within the continuous profile. Landmark annotation enables direct correlation between quantitative diameter changes and clinically recognized anatomical segments, facilitating interpretation across the full vessel length.

### High-Resolution Assessment of Aortic Aneurysm Morphology

Focused segment analysis of the aorta illustrates the ability of AortaPlot to characterize localized aneurysmal disease with high spatial resolution (Figure 4). Increased sampling density within the ascending aorta captures focal diameter expansion involving the sinuses of Valsalva and sinotubular junction. The landmark-annotated diameter profile delineates the extent and magnitude of aneurysmal dilation relative to adjacent segments, while standardized 3D renderings provide complementary visualization of root morphology across multiple viewing angles.

**Figure 4.**
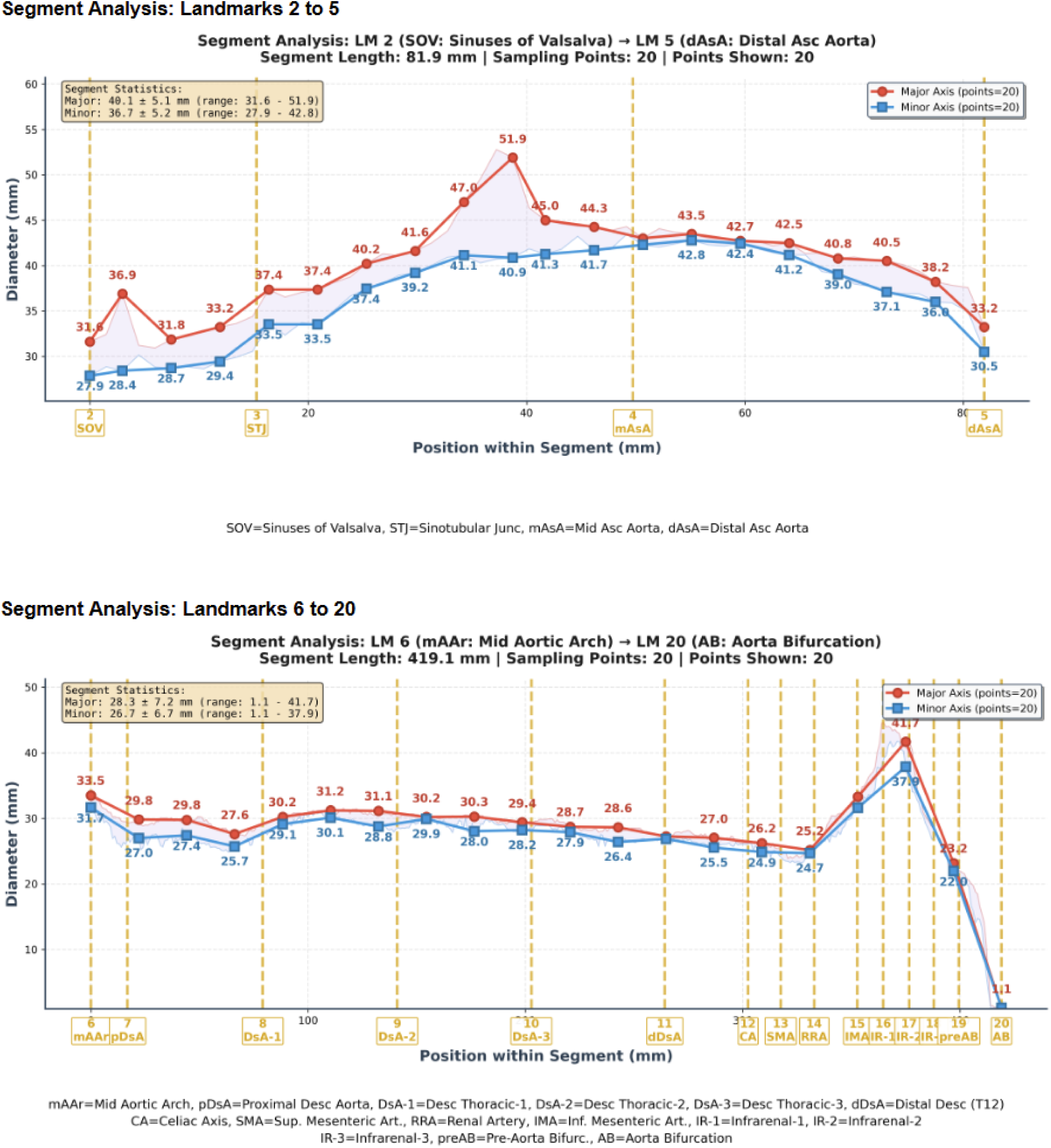
High-Resolution Segment Analysis of the Aorta Using Landmark-Guided Sampling. Focused quantitative analysis of the thoracic segment (Landmarks 2-5; top) and the abdominal segment (Landmarks 6-20; bottom) supports the assessment of thoracic aortic aneurysms and abdominal aortic aneurysms, respectively. Diameter measurements are sampled at uniformly spaced positions along the centerline, enabling high-resolution characterization of regional morphology. Major and minor diameters are shown for each sampling location to illustrate segment-specific variation and local maxima. Vertical dashed lines indicate the anatomical landmark boundaries (e.g., SOV, STJ, CA, AB) defining the analyzed regions. This framework facilitates a detailed evaluation of focal dilation patterns across both the thoracic and abdominal aorta.

### Quantitative Characterization of Aortic Aneurysm Extent

Analysis of an aortic aneurysm extending beyond the root demonstrates the utility of whole-vessel profiling for characterizing disease extent and segmental involvement (Figure 4). Continuous centerline-based measurements identify regions of maximal dilation and define transitions between aneurysmal and non-aneurysmal segments. The combined two-dimensional diameter profile and multi-angle 3D visualization enable comprehensive assessment of aneurysm morphology and spatial distribution within a single standardized report.

### Longitudinal Aortic Surveillance Using Continuous Centerline-Based Profiling

Application of AortaPlot to serial imaging studies demonstrates its ability to support longitudinal surveillance of aortic morphology using anatomically co-registered centerline-based measurements. Continuous diameter-versus-length profiles derived from multiple time points are aligned using automatically detected anatomical landmarks, preserving spatial correspondence across studies. Overlaid profiles enable direct visualization of interval changes in vessel diameter along the full length of the analyzed segment, facilitating identification of progressive dilation, regional remodeling, or stability over time (Figure 5). This landmark-guided longitudinal framework allows reproducible comparison across serial scans without reliance on manually selected axial measurement planes.

**Figure 5.**
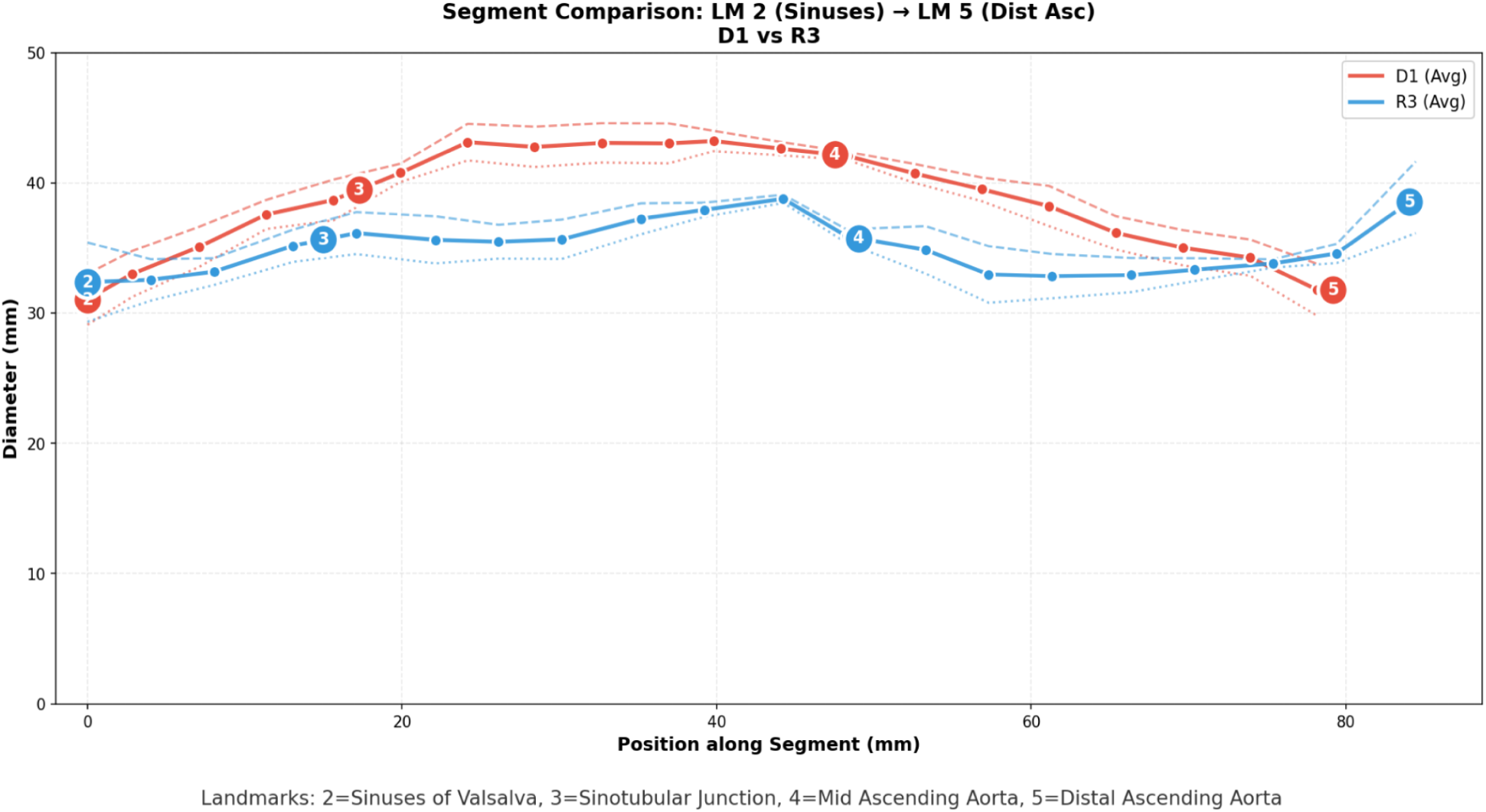
Longitudinal Aortic Diameter Comparison Across Landmarks Using Continuous Centerline-Based Profiling. Overlaid continuous diameter-versus-length profiles demonstrate longitudinal comparison of aortic morphology across the segment spanning Landmark 2 (Sinuses of Valsalva) to Landmark 5 (Distal Ascending Aorta). Profiles from two imaging time points are shown, enabling direct pointwise comparison along a consistent centerline parameterization. Anatomical landmark alignment preserves spatial correspondence across studies, facilitating assessment of interval changes in vessel diameter, regional progression or regression, and segment-specific remodeling. This longitudinal framework supports reproducible surveillance of aortic disease within standardized anatomical boundaries.

## Discussion

This study demonstrates the feasibility of a unified computational framework for reproducible, centerline-based quantification of aortic morphology across the full vessel length. AortaPlot enables continuous diameter assessment from the aortic root to the bifurcation, supports high-resolution regional sampling, and produces consistent two-dimensional and 3D representations suitable for clinical interpretation and longitudinal surveillance.

Across representative cases, AortaPlot reliably reconstructed full-length centerlines and generated orthogonal cross-sectional measurements using a configurable Sliding Axis approach. Continuous profiling captured physiologic tapering in normal anatomy and focal dilation in pathological cases, while regional oversampling enabled granular characterization of aneurysmal segments. Radiomic feature extraction was included to demonstrate feasibility of deriving standardized quantitative descriptors from anatomically ordered representations for machine learning and longitudinal modeling, rather than to evaluate predictive performance in this study. Accordingly, this manuscript intentionally focuses on representation and standardization as prerequisites for reliable downstream modeling, rather than advancing outcome prediction or performance claims.

Beyond cross-sectional assessment, the protocol supports reproducible longitudinal surveillance through consistent centerline parameterization and landmark-based alignment. Overlaid profiles enable pointwise comparison across time, addressing limitations of conventional workflows that rely on isolated axial measurements subject to inter-study variability.

The modular architecture formalizes segmentation, measurement, and visualization within a unified pipeline while allowing individual components to evolve independently. This design supports reproducibility across datasets, institutions, and emerging computational methods, while maintaining interpretability within clinical workflows.

AortaPlot establishes a reproducible, end-to-end computational framework for continuous, centerline-based characterization of aortic morphology from routine CT imaging. By standardizing segmentation, anatomical ordering, cross-sectional measurement, and landmark-annotated representation within a modular pipeline, this protocol addresses key sources of variability that limit comparability across datasets, institutions, and time points. Importantly, AortaPlot produces structured, anatomically contextualized outputs that are directly amenable to machine learning, radiomics, and longitudinal modeling, while remaining interpretable within clinical workflows.

While AortaPlot is a fully standalone system, its primary contribution lies in establishing a standardized, anatomically ordered representation of the aorta that enables consistent longitudinal analysis and supports advanced quantitative modeling. This combination of methodological rigor, modular extensibility, and standardized representation positions AortaPlot as a scalable foundation for multi-center validation, outcome-linked modeling, and future AI systems for aortic disease assessment.

### Limitations and Future Directions

The AortaPlot framework faces specific geometric challenges at the vascular terminals. Proximally, aortic root measurements (Landmarks 1-3) rely on standardized longitudinal offsets rather than discrete biological landmarks like the valve nadirs. These proxies may not fully capture asymmetric remodeling in severe root pathology or Type A dissections. Distally, the bifurcation (Landmark 20) is susceptible to “endpoint jitter” during skeletonization.

Future iterations will incorporate multi-class segmentation of the aortic valve and coronary ostia. Transitioning from geometric proxies to feature-based anatomical anchors will evolve the protocol from a surveillance tool into a high-precision platform for surgical-grade valvular planning.

## Data Availability

All data used in the present study are openly available online from the Aortic Vessel Tree (AVT) Collection and the AMOS Multi-Organ Benchmark. The reports and analysis outputs generated from these data are available from the authors upon reasonable request.

---

**Figure S1.**
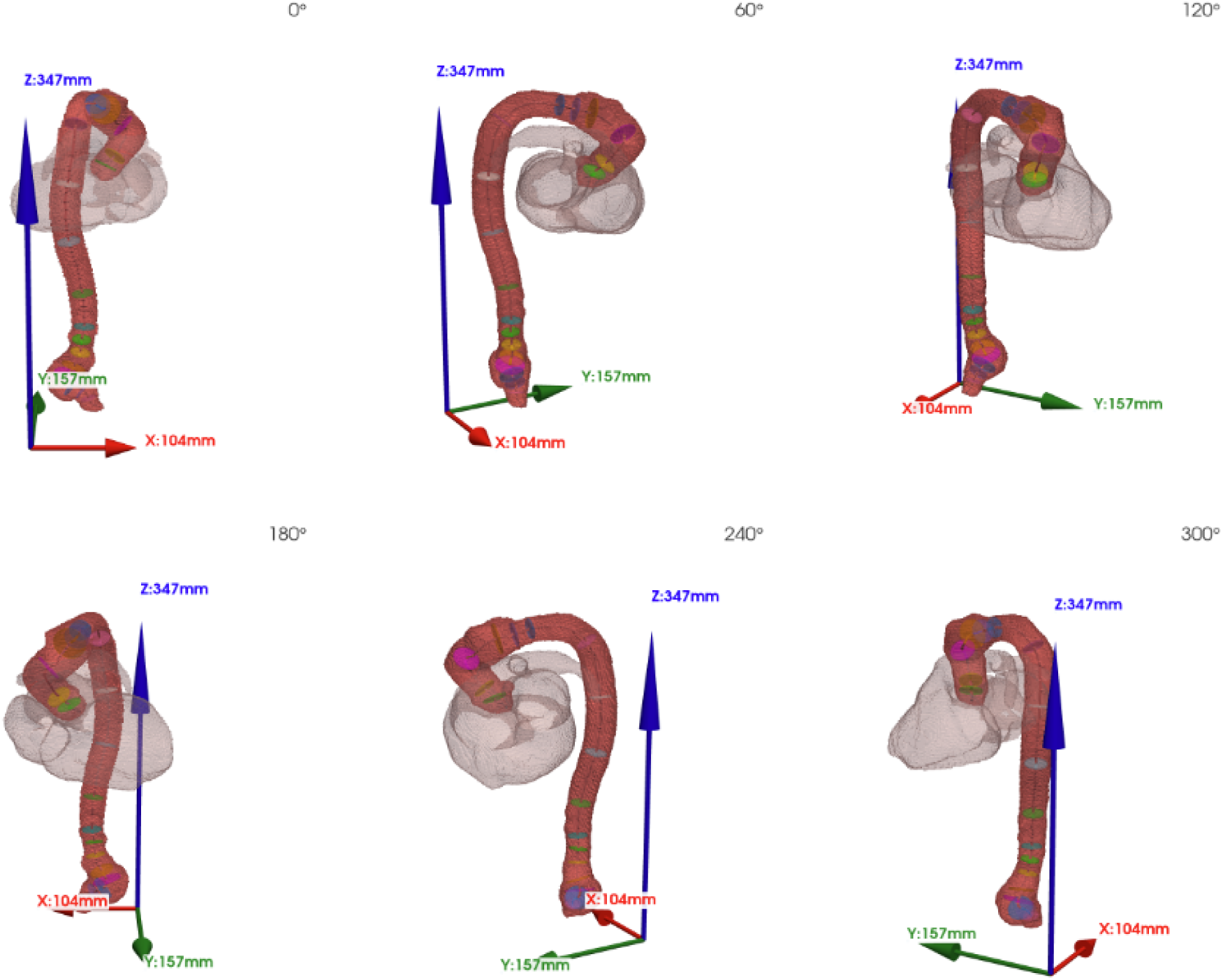
Standardized Multi-Angle 3D Visualization of an Aorta With an Abdominal Aortic Aneurysm Generated by AortaPlot.

**Table S1.** Definition and detection of standardized aortic anatomical landmarks. Standardized anatomical landmarks used for aortic centerline analysis and their corresponding detection rules. Landmark locations are determined using a hybrid approach that integrates anatomical reference masks from TotalSegmentator with geometric rules based on fixed offsets or relative centerline positions. When anatomical reference structures are unavailable or unreliable, predefined geometric offsets are applied to ensure consistent landmark placement across cases.

| Landmark | Anatomical Location | Vertebral Level | Detection Method | Abbreviation |
| --- | --- | --- | --- | --- |
| 1 | Annulus |  | Proximal centerline terminus (0% of centerline length) | ANN |
| 2 | Sinuses of Valsalva |  | Fixed offset 15 mm distal to annulus | SOV |
| 3 | Sinotubular Junction |  | Fixed offset 30 mm distal to annulus | STJ |
| 4 | Mid Ascending Aorta |  | Geometric midpoint between landmarks 3 and 5 | mAsA |
| 5 | Distal Ascending Aorta |  | Level of brachiocephalic trunk origin | dAsA |
| 6 | Mid Aortic Arch |  | Geometric midpoint between landmarks 5 and 7 | mAAr |
| 7 | Proximal Descending Aorta |  | Immediately distal to left subclavian artery origin | pDsA |
| 8 | Descending Thoracic Aorta-1 |  | 25% between landmarks 7 and 11 | DsA-1 |
| 9 | Descending Thoracic Aorta-2 |  | 50% between landmarks 7 and 11 | DsA-2 |
| 10 | Descending Thoracic Aorta-3 |  | 75% between landmarks 7 and 11 | DsA-3 |
| 11 | Distal Descending Aorta | T12 | Center of T12 vertebra | dDsA |
| 12 | Celiac Axis | T12-L1 | Midpoint between the vertebrae T12 and L1 (vertebral surrogate) | CA |
| 13 | Superior Mesenteric Artery | L1 | Center of L1 vertebral body | SMA |
| 14 | Right Renal Artery | L1-L2 | Midpoint between the vertebrae L1 and L2 | RRA |
| 15 | Inferior Mesenteric Artery | L3 | Center of L3 vertebral body | IMA |
| 16 | Infrarenal-1 |  | 25% of centerline distance between landmarks 15 and 20 | IR-1 |
| 17 | Infrarenal-2 |  | 50% of centerline distance between landmarks 15 and 20 | IR-2 |
| 18 | Infrarenal-3 |  | 75% of centerline distance between landmarks 15 and 20 | IR-3 |
| 19 | Pre-Aorta Bifurcation |  | Fixed offset 20 mm proximal to aortic bifurcation along centerline | preAB |
| 20 | Aorta Bifurcation |  | Distal centerline terminus at bifurcation into common iliac arteries (100% of centerline length) | AB |

## References

1. Fogante M, Esposto Pirani P, Cela F, Alfonsi J, Tagliati C, Balardi L, Argalia G, Di Eusanio M, Schicchi N. Computed tomography imaging of thoracic aortic surgery: Distinguishing life-saving repairs from life-threatening complications. J Imaging. 2025;11: 119.

2. Ricco J-B, Hostalrich A, Chaufour X. Aorta unveiled: the crucial role of imaging in diagnosing and managing aortic disease-a review. Eur Heart J Imaging Methods Pract. 2025;3: qyaf108.

3. Alexander KC, Ikonomidis JS, Akerman AW. New directions in diagnostics for aortic aneurysms: Biomarkers and machine learning. J Clin Med. 2024;13: 818.

4. Di Bella C, Coates PT, Mukherjee P, Visser J, Yue Z, editors. Bioengineering Solutions in Surgery: Advances, applications and solutions for clinical translation. Frontiers Media SA; 2022.

5. Wirth W, Eckstein F, Kemnitz J, Baumgartner CF, Konukoglu E, Fuerst D, Chaudhari AS. Accuracy and longitudinal reproducibility of quantitative femorotibial cartilage measures derived from automated U-Net-based segmentation of two different MRI contrasts: data from the osteoarthritis initiative healthy reference cohort. MAGMA. 2021;34: 337–354.

6. Jin X, Li Y, Yan F, Liu Y, Zhang X, Li T, Yang L, Chen H. Automatic coronary plaque detection, classification, and stenosis grading using deep learning and radiomics on computed tomography angiography images: a multi-center multi-vendor study. Eur Radiol. 2022;32: 5276–5286.

7. Zhang X, Zhang Y, Zhang G, Qiu X, Tan W, Yin X, Liao L. Deep learning with radiomics for disease diagnosis and treatment: Challenges and potential. Front Oncol. 2022;12: 773840.

8. Levenson RM, Singh Y, Rieck B, Hathaway QA, Farrelly C, Rozenblit J, Prasanna P, Erickson B, Choudhary A, Carlsson G, Sarkar D. Advancing precision medicine: Algebraic topology and differential geometry in radiology and computational pathology. Lab Invest. 2024;104: 102060.

9. Radl L, Jin Y, Pepe A, Li J, Gsaxner C, Zhao F-H, Egger J. AVT: Multicenter aortic vessel tree CTA dataset collection with ground truth segmentation masks. Data Brief. 2022;40: 107801.

10. Ji Y, Bai H, Yang J, Ge C, Zhu Y, Zhang R, Li Z, Zhang L, Ma W, Wan X, Luo P. AMOS: A large-scale abdominal multi-organ benchmark for versatile medical image segmentation. arXiv [eess.IV]. 2022. Available: http://arxiv.org/abs/2206.08023

11. Wasserthal J, Breit H-C, Meyer MT, Pradella M, Hinck D, Sauter AW, Heye T, Boll DT, Cyriac J, Yang S, Bach M, Segeroth M. TotalSegmentator: Robust segmentation of 104 anatomic structures in CT images. Radiol Artif Intell. 2023;5: e230024.

12. Isensee F, Jaeger PF, Kohl SAA, Petersen J, Maier-Hein KH. nnU-Net: a self-configuring method for deep learning-based biomedical image segmentation. Nat Methods. 2021;18: 203–211.

13. Gulati A, Zamirpour S, Leach J, Khan A, Wang Z, Xuan Y, Hope MD, Saloner DA, Guccione JM, Ge L, Tseng EE. Ascending thoracic aortic aneurysm elongation occurs in parallel with dilatation in a nonsurgical population. Eur J Cardiothorac Surg. 2023;63. doi:10.1093/ejcts/ezad241

14. Accarino G, Oliva A, Buongiorno F, Cantile N, Pollio C, DI Bonito C, Bracale UM, DE Vries J-PPM. Association of longitudinal aortoiliac morphometric changes after endovascular aneurysm repair with outcomes. J Cardiovasc Surg (Torino). 2025. doi:10.23736/S0021-9509.25.13422-8

